# Diphtheria resurgence, drug-supply delays, and case fatality at a tertiary hospital in Adamawa State, north-eastern Nigeria: a retrospective cohort study (2023–2026)

**DOI:** 10.64898/2026.07.07.26357438

**Authors:** Hayatu Ahmed, Ahmad Hayatu

## Abstract

**Background:** Diphtheria caused by toxigenic *Corynebacterium diphtheriae* re-emerged in Nigeria from December 2022 as the country’s largest ever recorded outbreak, accumulating over 20,000 suspected cases and 872 confirmed deaths by December 2025. The Borno-Adamawa-Yobe (BAY) states zone in north-eastern Nigeria is a recognised high-vulnerability cluster, yet no facility-level, longitudinal epidemiological data from Adamawa State have been published.

**Methodology/Principal findings:** We conducted a retrospective cohort study of all patients admitted with diphtheria to the Isolation Ward of Modibbo Adama University Teaching Hospital (MAUTH), Yola — the sole federal university teaching hospital in Adamawa State — from January 2023 to April 2026, using prospective admission register records. Sixty-one patients were identified (17.9% of 330 total isolation admissions). Admissions escalated 580% from 5 (2023) to 34 (2025). Median age was 8.0 years; 91.8% were under 15 years. The overall in-hospital case fatality rate (CFR) was 41.5% (22/53 known outcomes; 95% confidence interval: 29.0–55.0%). Annual CFR declined from 60.0% (2023) to 28.6% (2025), temporally consistent with improving diphtheria antitoxin access. A critical operational finding was the persistent shortage of intravenous erythromycin — the mandated antibiotic for patients unable to swallow — compelling oral administration in patients with pharyngeal pseudomembrane and dysphagia. Respiratory distress at presentation carried an 80% CFR; cardiac complication, 100%. Age and sex were not statistically significant mortality predictors. Gombi local government area contributed 16.4% of cases — the highest burden among non-capital communities — consistent with its role as a population movement corridor from Borno State’s outbreak epicentre. A September–October seasonal peak (47.5% of admissions) was identified, diverging from the national January–April pattern.

**Conclusions/Significance:** This study provides the first peer-reviewed, facility-level diphtheria epidemiological dataset from Adamawa State. The in-hospital CFR substantially exceeds national surveillance averages due to referral bias and historical drug supply constraints. The declining CFR against rising admissions signals improving case management. Pre-positioning of diphtheria antitoxin and intravenous erythromycin before each August–October peak, accelerated childhood immunisation catch-up, and strengthened surveillance in Adamawa State are identified as urgent priorities.

**Author summary:** Diphtheria is a throat infection caused by bacteria that produce a toxin capable of damaging the heart and nerves. It is preventable by childhood vaccination, but large outbreaks occur when vaccination rates fall. Nigeria is currently experiencing its worst recorded diphtheria outbreak, with over 20,000 cases and nearly 900 deaths since late 2022.

We studied all diphtheria patients admitted to Modibbo Adama University Teaching Hospital in Yola, north-eastern Nigeria — the region’s only federal teaching hospital — over three and a half years. We found that admissions more than tripled in that period, nearly 9 in 10 patients were children under 15, and over 4 in 10 who had a documented outcome died in hospital. One of our most important findings was a shortage of intravenous antibiotic medicine: patients with severe throat infection cannot swallow tablets, but the injectable form was repeatedly unavailable, forcing the team to crush tablets or use stomach tubes in critically ill children — a dangerous workaround.

We identified a distinctive peak in September and October, and a rural cluster in Gombi that appears to receive patients from the neighbouring state with the highest national burden. Our study highlights that reliable supply of the correct antibiotic formulation, continued vaccination campaigns, and advance planning for the seasonal peak could save children’s lives in this under-resourced region.

## Introduction

*Corynebacterium diphtheriae*, the causative agent of diphtheria, produces a potent exotoxin responsible for the disease’s most life-threatening manifestations — upper airway obstruction from pharyngeal pseudomembrane extension, myocarditis, and demyelinating polyneuropathy [1]. Although diphtheria is entirely preventable through diphtheria-tetanus-pertussis (DTP) vaccination, global resurgence has followed the erosion of herd immunity in populations with chronically suboptimal vaccine coverage [2]. Case fatality rate (CFR) ranges from 5–10% when diphtheria antitoxin (DAT) and appropriate antibiotics are available, but may reach 20–40% in resource-limited settings where access to these treatments is intermittent or absent [3].

Nigeria’s current diphtheria epidemic — the largest in the country’s recorded surveillance history — began in Kano State in December 2022. By December 2025, the Nigeria Centre for Disease Control and Prevention (NCDC) had documented over 20,000 suspected cases, 15,273 confirmed cases, and 872 confirmed-case deaths nationally [4,5]. The epidemic has been characterised by overwhelming predominance in children aged 1–14 years, near-universal absence of prior vaccination among confirmed cases, and progressive geographic spread from the north-western epicentre to the north-eastern states [6]. Crucially, antimicrobial susceptibility testing of Nigerian *C. diphtheriae* isolates from May 2022 to July 2023 demonstrated complete resistance to all penicillins while retaining erythromycin susceptibility — a finding that established erythromycin (or azithromycin) as the mandatory antibiotic of choice and rendered existing penicillin stocks clinically useless [7]. The 2024 WHO Clinical Management of Diphtheria Guideline formalised this as a strong recommendation [8].

Within the Borno-Adamawa-Yobe (BAY) States zone — a WHO-designated humanitarian emergency cluster — the World Health Organization documented 417 suspected diphtheria cases and 16 deaths (CFR 3.8%) during the first eight epidemiological weeks of 2024, with Adamawa State officially contributing only 2 cases versus Borno State’s 271 [9]. This striking undercount, relative to facility-based capture, illustrates the structural limitations of passive notification surveillance and the critical complementary role of hospital-based epidemiology. In 2018, Adamawa State was one of four states reporting Nigeria’s largest pre-current-epidemic diphtheria outbreak [10]. Modibbo Adama University Teaching Hospital (MAUTH), Yola — the sole federal university teaching hospital in Adamawa State, serving an estimated catchment population of 4.3 million — functions as the principal tertiary referral centre for infectious disease management in the region.

Despite Adamawa State’s documented vulnerability, no peer-reviewed, facility-level, longitudinal epidemiological analysis of diphtheria has been published from this region. This study addresses that evidence gap through a retrospective cohort analysis of 61 diphtheria admissions at MAUTH from January 2023 to April 2026, with the specific aims of: (i) describing demographic profile, temporal trends, and geographic distribution of admissions; (ii) documenting complication-specific CFRs and the clinical impact of intravenous erythromycin shortage; (iii) characterising annual CFR trends in relation to antitoxin access; and (iv) contextualising findings against national, north-eastern regional, and international benchmarks to inform evidence-based interventions in Adamawa State.

## Methods

### Study design and setting

A hospital-based retrospective cohort study was conducted at the Isolation Ward of MAUTH, Yola, Adamawa State, Nigeria — a 350-bed federal tertiary institution and the only federal university teaching hospital in Adamawa State. The isolation ward manages patients with suspected or confirmed notifiable infectious diseases including COVID-19, viral haemorrhagic fever (VHF)/Lassa Fever, diphtheria, and monkeypox.

### Ethics statement

This study was approved by the Health Research Ethics Committee of Modibbo Adama University Teaching Hospital, Yola, Adamawa State, Nigeria (MAUTHYOLA/HREC/26/437), dated 26th March 2026. The committee waived the requirement for individual informed consent because the study used routinely collected, de-identified records. The work was conducted in accordance with the Declaration of Helsinki.

### Data source and study period

#### S1 Dataset. De-identified analysis-ready dataset and variable codebook

De-identified, analysis-ready dataset (330 records from the MAUTH Isolation Ward Register, 2020–2026, of which 61 are diphtheria admissions analysed in this study) provided in XLSX format, together with a variable codebook defining each field. Fields include: sequential case identifier; year and month of admission; sex; age group; under-15 status; clinical presentation category; local government area cluster; outcome category; mortality indicator; and length of stay (days). No patient names, exact dates of birth, or individual identifying information are included. The ethics approval and de-identification statement are included in the dataset documentation sheet. The analysis code is available from the corresponding author on reasonable request.

### Inclusion and exclusion criteria

All patients admitted with a primary diagnosis of suspected or confirmed diphtheria were included. Patients admitted for other conditions without a diphtheria differential diagnosis were excluded.

### Case definitions

Suspected diphtheria was defined as an acute upper respiratory illness with pharyngeal, tonsillar, or laryngeal pseudomembrane, consistent with NCDC case definitions. Confirmed diphtheria required laboratory confirmation via *C. diphtheriae* culture from throat swab, or clinical compatibility plus epidemiological linkage to a confirmed case, per NCDC and WHO definitions [3,11].

### Variables and measurements

Variables extracted from the register included: age (years); sex; dates of admission and discharge or death; clinical diagnosis text; patient local government area (LGA) of origin; and outcome (Death; Discharged; Transferred; Left Against Medical Advice; Unknown). Length of stay (LOS) was calculated as discharge or death date minus admission date. Clinical presentation categories were derived from recorded diagnosis text using standardised keyword mapping. Vaccination history and antibiotic formulation (oral versus intravenous) were recorded where documented in the register. Intravenous erythromycin availability at MAUTH during each period was ascertained through clinical recall and pharmacy records.

### Statistical analysis

Data were analysed using Python 3.12 with NumPy 1.26, Pandas 2.2, SciPy 1.17, and Matplotlib 3.8. CFR was calculated as deaths divided by known outcomes, multiplied by 100. Wilson score 95% confidence intervals (CIs) were computed for annual CFRs. Age was categorised as: <5, 5–9, 10–14, 15–19, 20–29, 30–44, and ≥45 years, with <5 years as the reference group. Crude odds ratios (ORs) with 95% CIs were calculated using the Woolf method. Chi-square and Fisher’s exact tests assessed categorical associations. Binary logistic regression (Broyden-Fletcher-Goldfarb-Shanno optimisation; Nagelkerke pseudo-R²) identified independent mortality predictors. LOS differences between groups were assessed using the Mann-Whitney U test. All tests were two-tailed with a significance threshold of α=0.05. Statistical significance was defined as p<0.05.

## Results

### Overview and annual trends

Of 330 total MAUTH isolation admissions from May 2020 to April 2026, diphtheria was the second most common diagnosis (n=61; 17.9%), after COVID-19 and community-acquired pneumonia (n=188; 57.0%) and before VHF/Lassa Fever (n=53; 16.1%), with no admissions documented before January 2023 (Table 1). Annual admissions escalated from 5 (2023) to 18 (2024) to 34 (2025) — a 580% increase in three years — with four further admissions in the first quarter of 2026 (Table 2 and Fig 3).

**Table 1.** MAUTH Isolation Ward admissions by disease category (2020–2026).

| Disease category | n | % of total | First recorded |
| --- | --- | --- | --- |
| COVID-19/community-acquired pneumonia | 188 | 57 | May-20 |
| Diphtheria | 61 | 17.9 | Jan-23 |
| VHF/Lassa Fever | 53 | 16.1 | Oct-20 |
| Monkeypox | 15 | 4.5 | 2022 |
| Other | 13 | 3.9 | Variable |
| Total | 330 | 100 | May-20 |

*VHF, viral haemorrhagic fever. Diphtheria admissions exclusively from January 2023, consistent with the national epidemic onset in December 2022.*

**Table 2.**
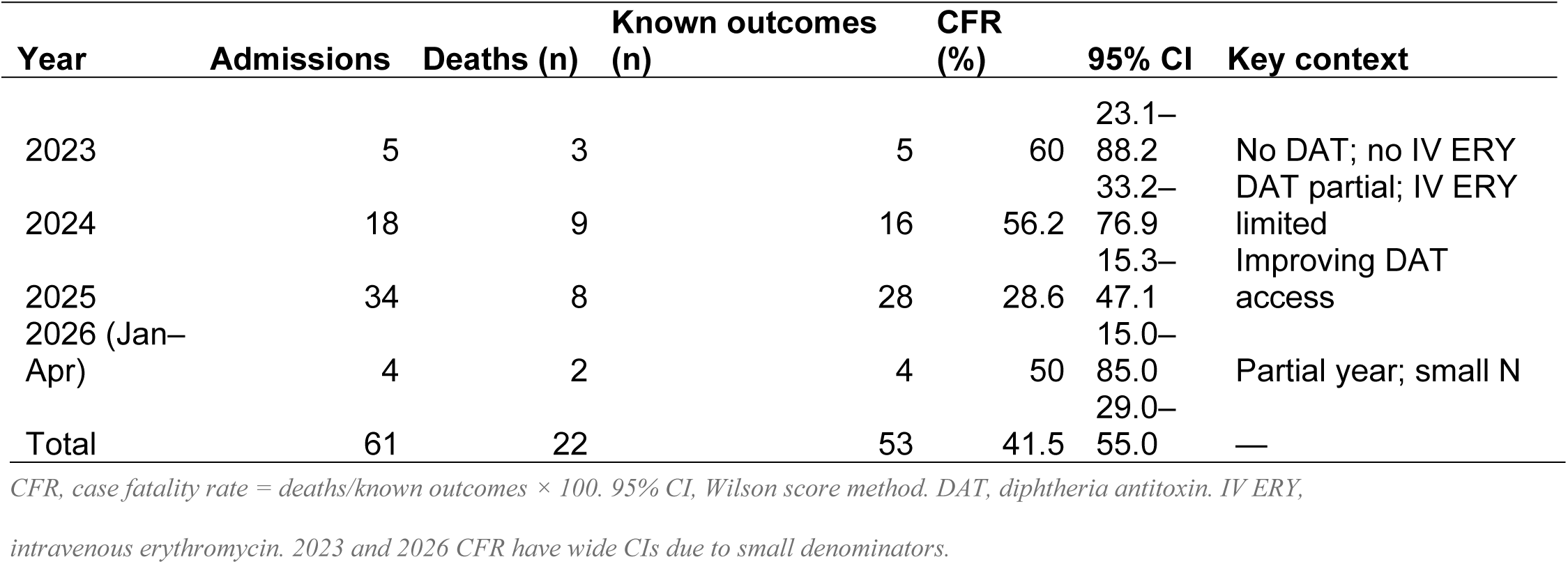
Annual diphtheria admissions, deaths, and case fatality rate with Wilson score 95% confidence intervals, MAUTH Yola (2023–2026).

Annual CFR declined progressively from 60.0% in 2023 (95% CI: 23.1–88.2%) to 56.2% in 2024 (95% CI: 33.2–76.9%) and 28.6% in 2025 (95% CI: 15.3–47.1%), temporally consistent with progressive procurement and distribution of DAT through NCDC–WHO–UNICEF from mid-2023 onwards (Table 2 and Fig 3) [12]. The 2023–2024 CFR decline also coincided with progressive improvement in intravenous erythromycin availability, albeit incomplete throughout the study period.

### Epidemic curve and seasonal pattern

The monthly epidemic curve (Fig 1) revealed a pronounced September–October seasonal peak, accounting for 29 of 61 admissions (47.5%) and 10 of 22 deaths aggregated across 2023–2026. This pattern diverges from the nationally reported January–April dry-season clustering observed in Kano-centred outbreak data [10]. March had the highest monthly CFR (100%; 3/3), reflecting the early epidemic period with absent DAT and intravenous erythromycin.

**Fig 1.**
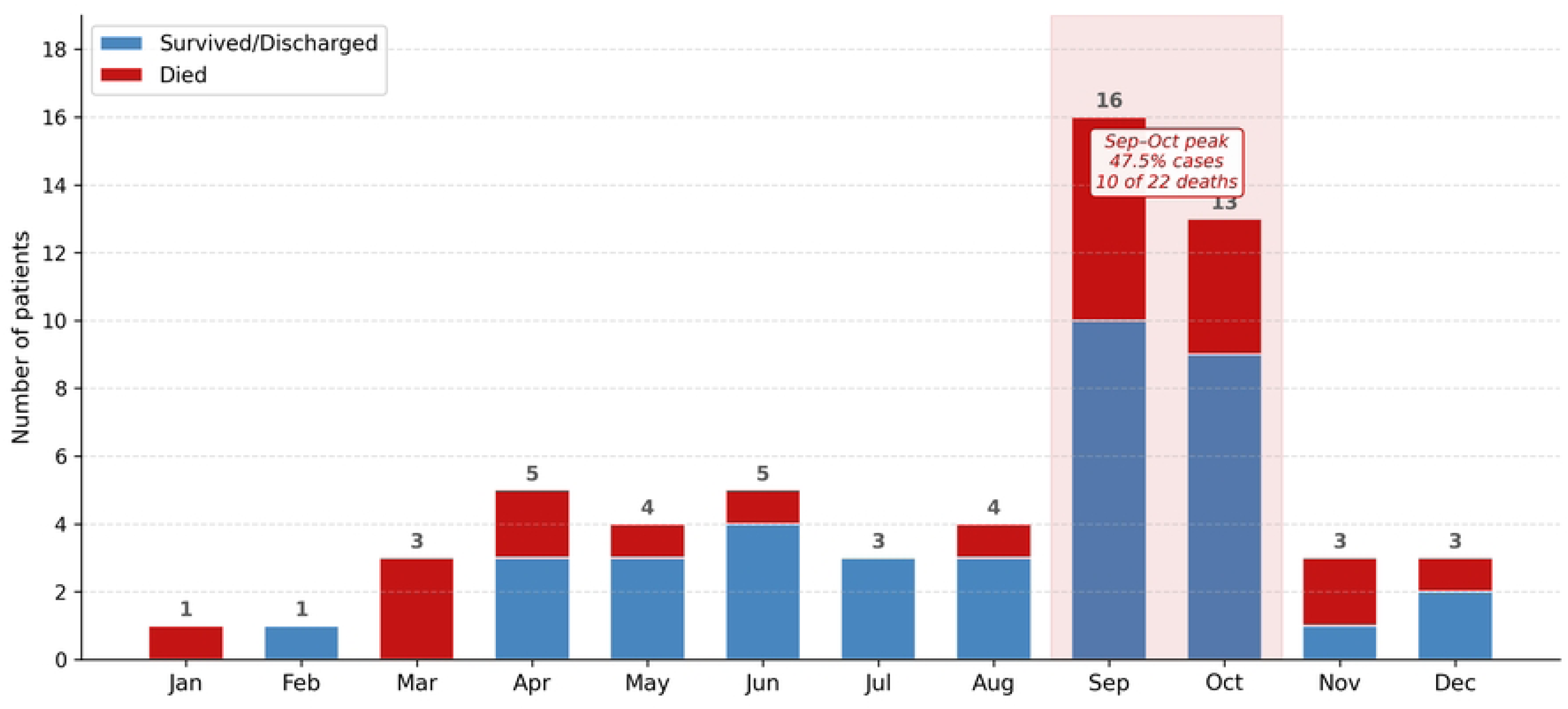
Monthly distribution of diphtheria admissions by outcome, MAUTH Yola, 2023–2026 (N=61). Blue bars, survived/discharged; red bars, died. September–October accounts for 47.5% of all admissions and 10 of 22 deaths. March 2023 CFR (100%; 3/3) reflects the early epidemic period with absent diphtheria antitoxin and intravenous erythromycin. CFR, case fatality rate; DAT, diphtheria antitoxin.

### Demographic characteristics

Twenty-eight males (45.9%) and 33 females (54.1%) were admitted. Ages ranged from 2 to 70 years (mean: 10.4 ± 10.0 years; median: 8.0 years; interquartile range [IQR]: 6.0–12.0). Children under 15 years constituted 91.8% (56/61) of admissions. The 5–9-year group was the largest stratum (n=27; 44.3%), followed by 10–14 years (n=11; 18.0%), <5 years (n=9; 14.8%), 15–19 years (n=5; 8.2%), and adults ≥20 years (n=9; 14.8%) (Table 3 and Fig 2).

**Fig 2.**
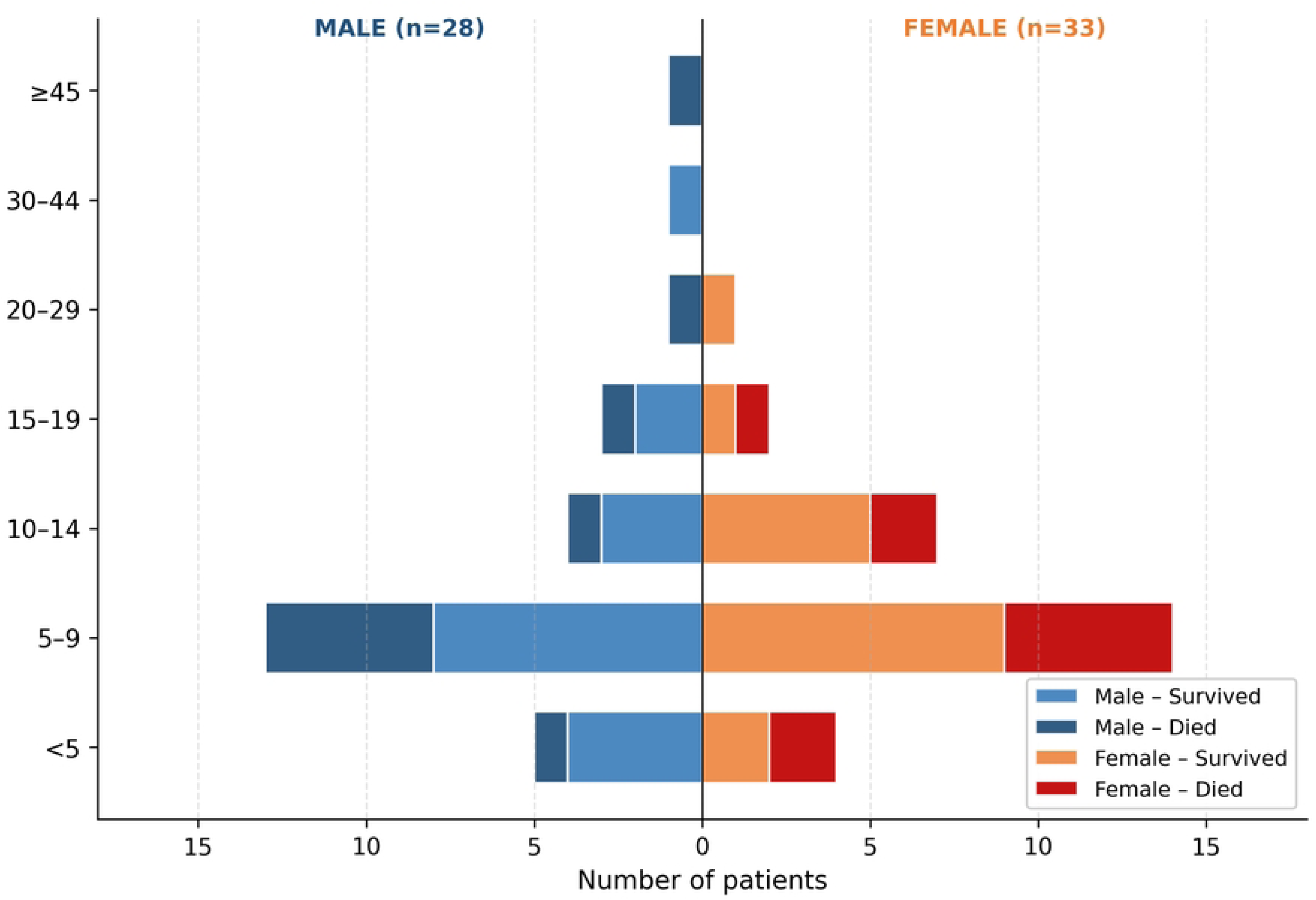
Age–sex population pyramid of diphtheria admissions, MAUTH Yola, 2023–2026 (N=61). Dark shading, died; light shading, survived. The 5–9-year group was the largest stratum (44.3%; n=27). Case fatality rates were uniformly high (33–57%) across all paediatric age groups, consistent with universal non-immune status in the admitted cohort. CFR, case fatality rate.

**Table 3.**
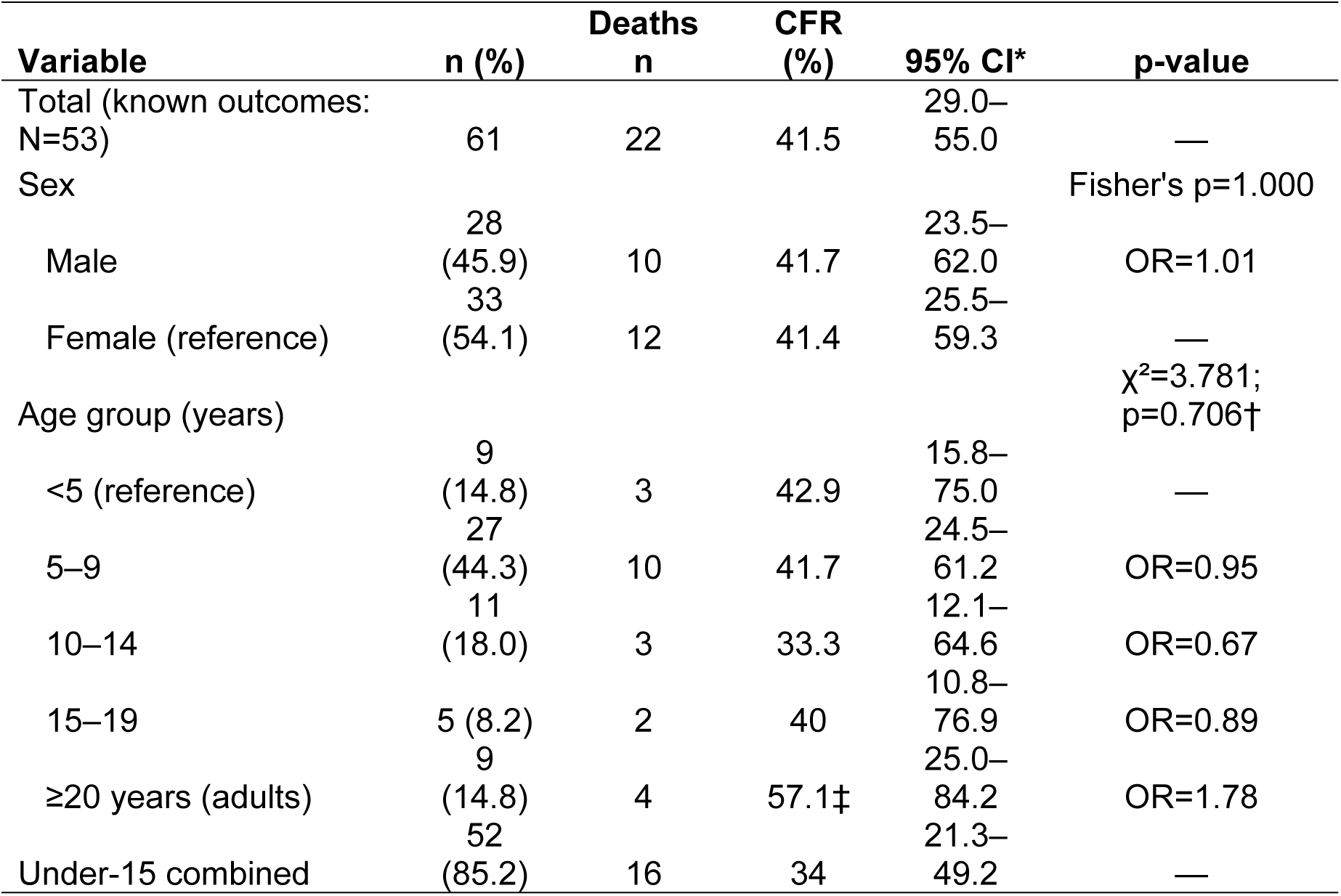
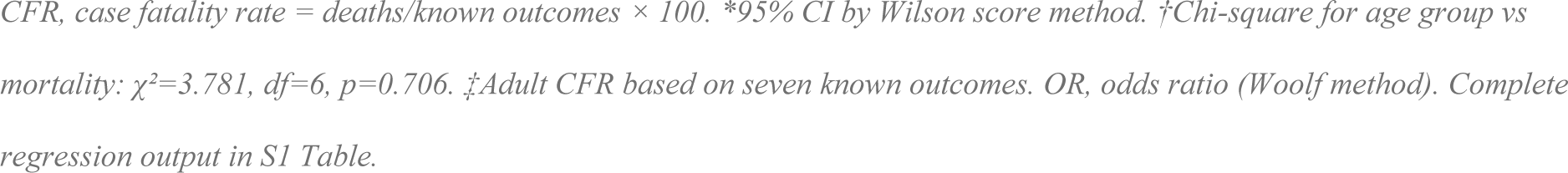
Sociodemographic characteristics and case fatality rates, MAUTH Yola diphtheria cohort (N=61).

| Variable | n (%) | Deaths<br>n | CFR<br>(%) | 95% CI* | p-value |
| --- | --- | --- | --- | --- | --- |
| Total (known outcomes:<br>N=53) | 61 | 22 | 41.5 | 29.0–<br>55.0 | — |
| Sex |  |  |  |  | Fisher's p=1.000 |
| Male | 28<br>(45.9) | 10 | 41.7 | 23.5–<br>62.0 | OR=1.01 |
| Female (reference) | 33<br>(54.1) | 12 | 41.4 | 25.5–<br>59.3 | — |
| Age group (years) | | | | | $\chi^2=3.781$;<br>p=0.706† |
| <5 (reference) | 9<br>(14.8) | 3 | 42.9 | 15.8–<br>75.0 | — |
| 5–9 | 27<br>(44.3) | 10 | 41.7 | 24.5–<br>61.2 | OR=0.95 |
| 10–14 | 11<br>(18.0) | 3 | 33.3 | 12.1–<br>64.6 | OR=0.67 |
| 15–19 | 5 (8.2) | 2 | 40 | 10.8–<br>76.9 | OR=0.89 |
| $\geq 20$ years (adults) | 9<br>(14.8) | 4 | 57.1‡ | 25.0–<br>84.2 | OR=1.78 |
| Under-15 combined | 52<br>(85.2) | 16 | 34 | 21.3–<br>49.2 | — |

CFR, case fatality rate = deaths/known outcomes × 100. \*95% CI by Wilson score method. †Chi-square for age group vs mortality: $\chi^2=3.781$ , $df=6$ , $p=0.706$ . ‡Adult CFR based on seven known outcomes. OR, odds ratio (Woolf method). Complete regression output in S1 Table.

### Clinical outcomes and case fatality

Of 61 admissions, 53 (86.9%) had documented outcomes: 22 died (CFR 41.5%); 21 survived or were transferred (34.4%); seven discharged confirmed positive (11.5%); three discharged confirmed negative (4.9%); and three left against medical advice (4.9%). Fig 3 shows outcome distribution and annual CFR trends.Sex-specific CFRs were 41.7% (male) and 41.4% (female); Fisher’s exact: OR=1.01, 95% CI: 0.33–3.11, p=1.000. Chi-square for age group versus mortality: χ²=3.781, df=6, p=0.706. Binary logistic regression (N=53 known outcomes; Nagelkerke pseudo-R²=0.127) identified no statistically significant independent predictor of mortality (complete regression output: S1 Table).

**Fig 3.**
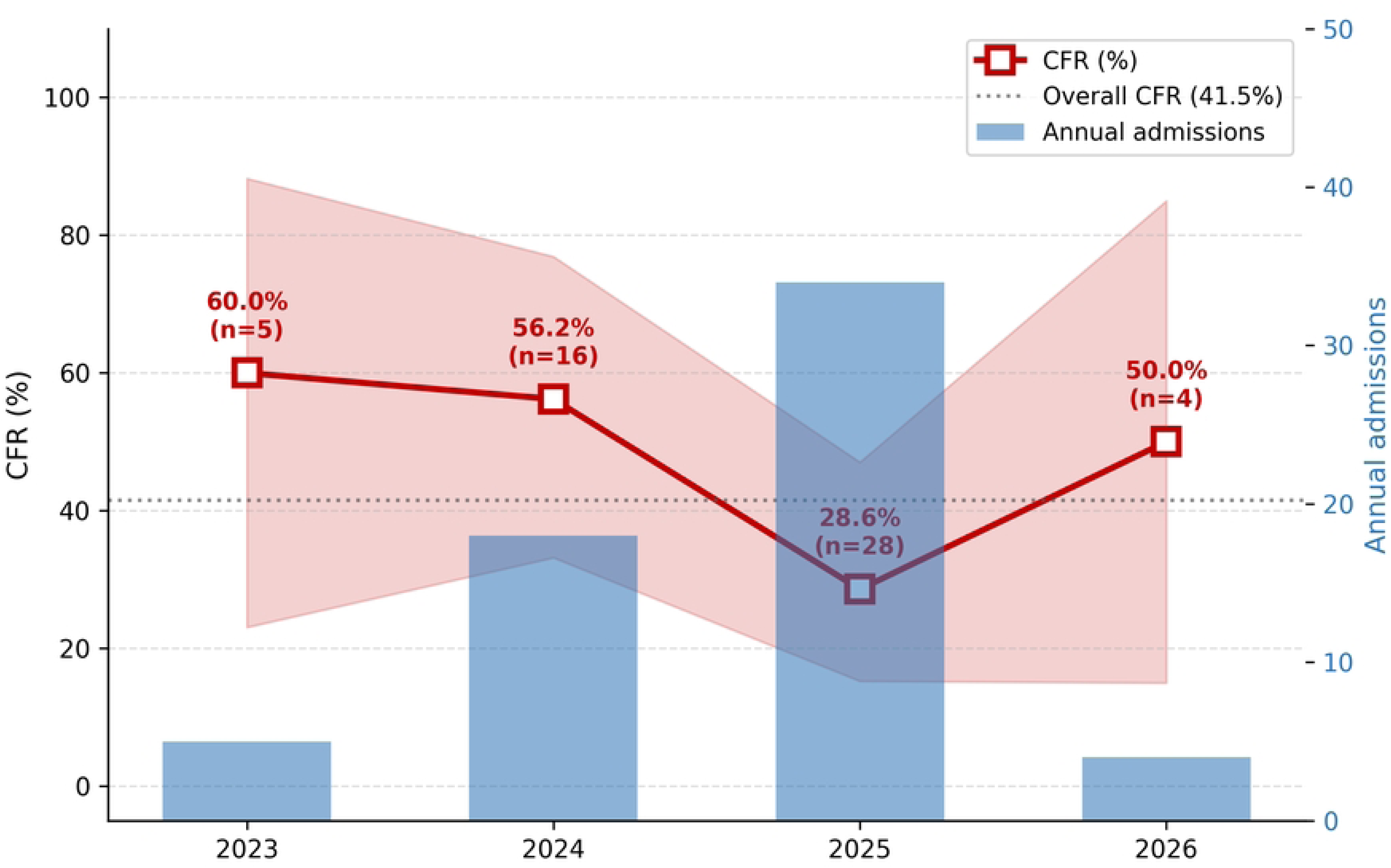
Annual diphtheria admissions and CFR with Wilson score 95% confidence intervals, MAUTH Yola, 2023–2026. Blue bars (right axis), annual admissions; red markers and shading, CFR with 95% CI. Admission escalation (580%; 5→34 cases) is shown alongside declining CFR (60.0%→28.6%), temporally consistent with progressive diphtheria antitoxin procurement from mid-2023 onwards. The 2026 CFR (50.0%) is based on four known outcomes and should be interpreted with caution. CFR, case fatality rate; CI, confidence interval; DAT, diphtheria antitoxin.

### Clinical presentation and complication-specific CFR

Suspected or unspecified pharyngeal diphtheria accounted for 77.0% of presentations (n=47; CFR 35.9%). Five patients presented with respiratory distress (CFR 80.0%), reflecting laryngeal pseudomembrane extension causing acute airway obstruction — the most immediately life-threatening diphtheria presentation [1]. One patient had a documented cardiac complication (CFR 100%), consistent with exotoxin-mediated myocarditis, which accounts for 50–60% of non-airway diphtheria deaths globally [1,8]. Peripheral neuropathy was documented in one patient who survived (CFR 0.0%) (Table 4 and Fig 4).

**Fig 4.**
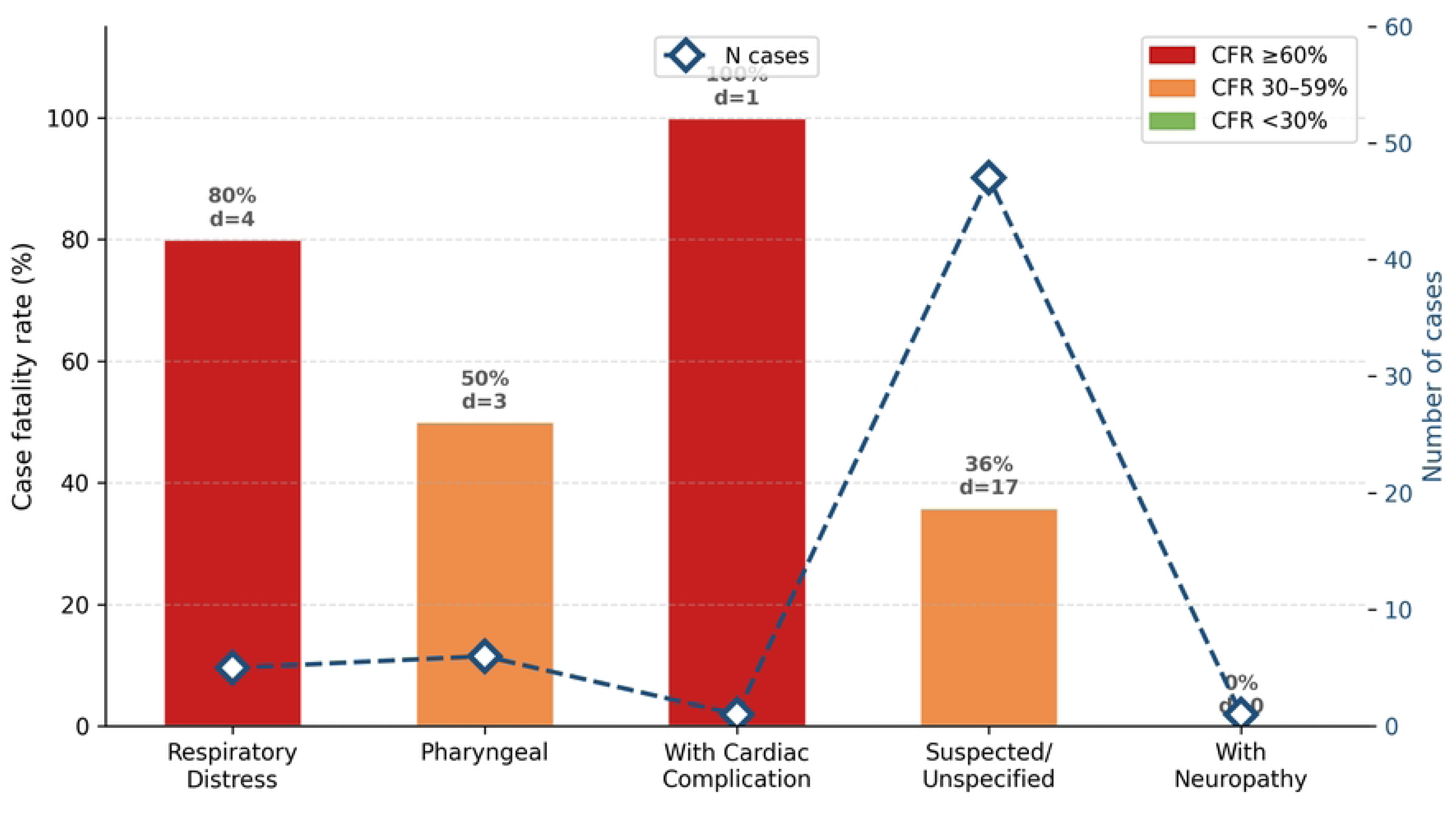
Case fatality rate (CFR) by clinical presentation category, MAUTH Yola diphtheria cohort, 2023–2026 (N=61). Bars show CFR by category. Diamonds (right axis) indicate case count. Double asterisks (**) denote CFR ≥50%. Respiratory distress (CFR 80.0%) and cardiac complication (CFR 100%) represent the highest-fatality presentations. CFR, case fatality rate; CCF, congestive cardiac failure.

**Table 4.**
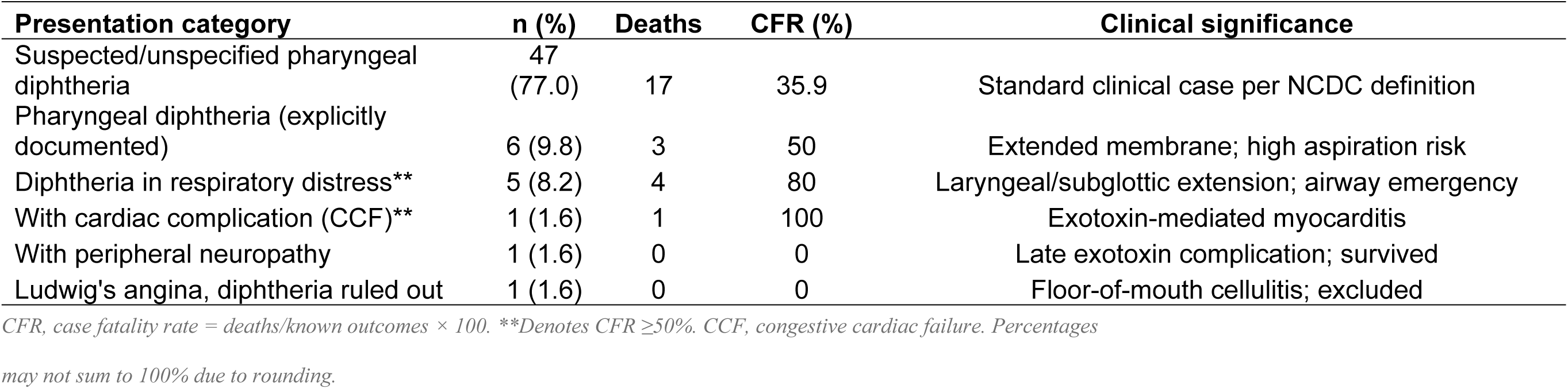
Clinical presentation categories and complication-specific case fatality rates, MAUTH Yola diphtheria cohort (N=61).

### Intravenous erythromycin shortage

A persistent shortage of intravenous (IV) erythromycin constituted a critical clinical challenge throughout the 2023–2026 study period. The 2024 WHO Clinical Management of Diphtheria Guideline recommends macrolide antibiotics — erythromycin or azithromycin — as a strong recommendation over penicillin, based on documented universal penicillin resistance in Nigerian isolates [8]. NCDC antimicrobial susceptibility data from May 2022 to July 2023 confirmed that all *C. diphtheriae* isolates tested were resistant to penicillin while retaining erythromycin susceptibility [7]. WHO and MSF guidelines specify that IV administration is mandatory when patients cannot swallow oral medication [8,13].

Pharyngeal diphtheria — the dominant presentation in this cohort — causes severe oropharyngeal dysphagia through pseudomembrane-mediated mechanical obstruction, acute mucosal pain, and cervical oedema [14]. In patients with laryngeal extension and respiratory distress (8.2% of this cohort; CFR 80%), oral drug administration risks aspiration into a critically narrowed airway. At MAUTH, the clinical team was compelled to administer oral erythromycin by crushing tablets or through nasogastric tubes in severely dysphagic patients — practices carrying risk of reduced bioavailability from acid degradation of crushed enteric-coated tablets, traumatic pseudomembrane dislodgement from nasogastric tube placement, and subtherapeutic plasma drug levels. This supply crisis was not unique to MAUTH: a prospective diphtheria treatment cohort from Yobe State Specialist Hospital Potiskum reported that “DAT and intravenous erythromycin, medications that are known to enhance favorable disease outcome, are not readily available” in north-eastern Nigeria, and documented a 100% increase in fatality rate with each 24-hour delay in appropriate treatment [15]. The WHO Grade 2 Emergency response procured 15,000 vials of IV erythromycin nationally by August 2023, but distribution constraints delayed access in Adamawa State relative to Kano and Yobe [12].

### Diphtheria antitoxin shortage and administration delays

Diphtheria antitoxin (DAT) — the only specific treatment neutralising circulating exotoxin and the single most important determinant of survival — was in critically short supply at MAUTH throughout the study period, mirroring shortages at subnational (Adamawa State) and national levels. DAT is not routinely stocked at Nigerian health facilities [21], and MAUTH held no independent standing DAT stock. Under the national DAT deployment protocol, the treating facility must request DAT through the Local Government Area and State Disease Surveillance and Notification Officer (DSNO) and State Epidemiologist, who escalate the requisition — supported by case investigation, laboratory, and adverse-event data — to the Diphtheria Emergency Operations Centre at the Nigeria Centre for Disease Control and Prevention (NCDC), where the National Incident Manager authorises release against eligibility criteria and available national stock [7,21].

This multi-tier requisition pathway introduced substantial delays between clinical diagnosis and DAT administration, even when DAT was ultimately available. Delays were most pronounced for patients presenting during weekends and outside routine working hours, when the DSNO reporting and national release chain operated with reduced availability. In several instances, patients with clinically advanced, toxin-mediated disease deteriorated — progressing to respiratory compromise, myocarditis, or death — before DAT could be obtained and administered. Because DAT efficacy is critically time-dependent, with maximal benefit only when given early in the disease course [1], these administrative and out-of-hours delays represented a modifiable contributor to the high early case fatality observed in this cohort (median length of stay among those who died was 3 days; Table 2 and S2 Table), compounding the parallel shortage of intravenous erythromycin described above.

### Length of stay and geographic distribution

Of 61 admissions, 44 (72.1%) had documented admission and discharge or death dates. Median LOS was 6 days (IQR: 2–31; mean: 18.4 ± 27.1 days; range: 0–90). Patients who died had a shorter median LOS (3 days; mean 9.7 days) than survivors (median 6 days; mean 24.6 days), consistent with early-admission death from advanced disease (Mann-Whitney U=177, p=0.139; S2 Table). Geographic analysis identified Yola South (n=17; 27.9%), Yola North and Jimeta (n=11; 18.0%), and Gombi LGA (n=10; 16.4%) as the three largest contributor communities (Fig 5 and Fig 6). Gombi — a rural market centre proximal to Borno State’s outbreak epicentre with documented population movement from conflict-affected internally displaced person (IDP) communities — was the highest-burden non-capital cluster.

**Fig 5.**
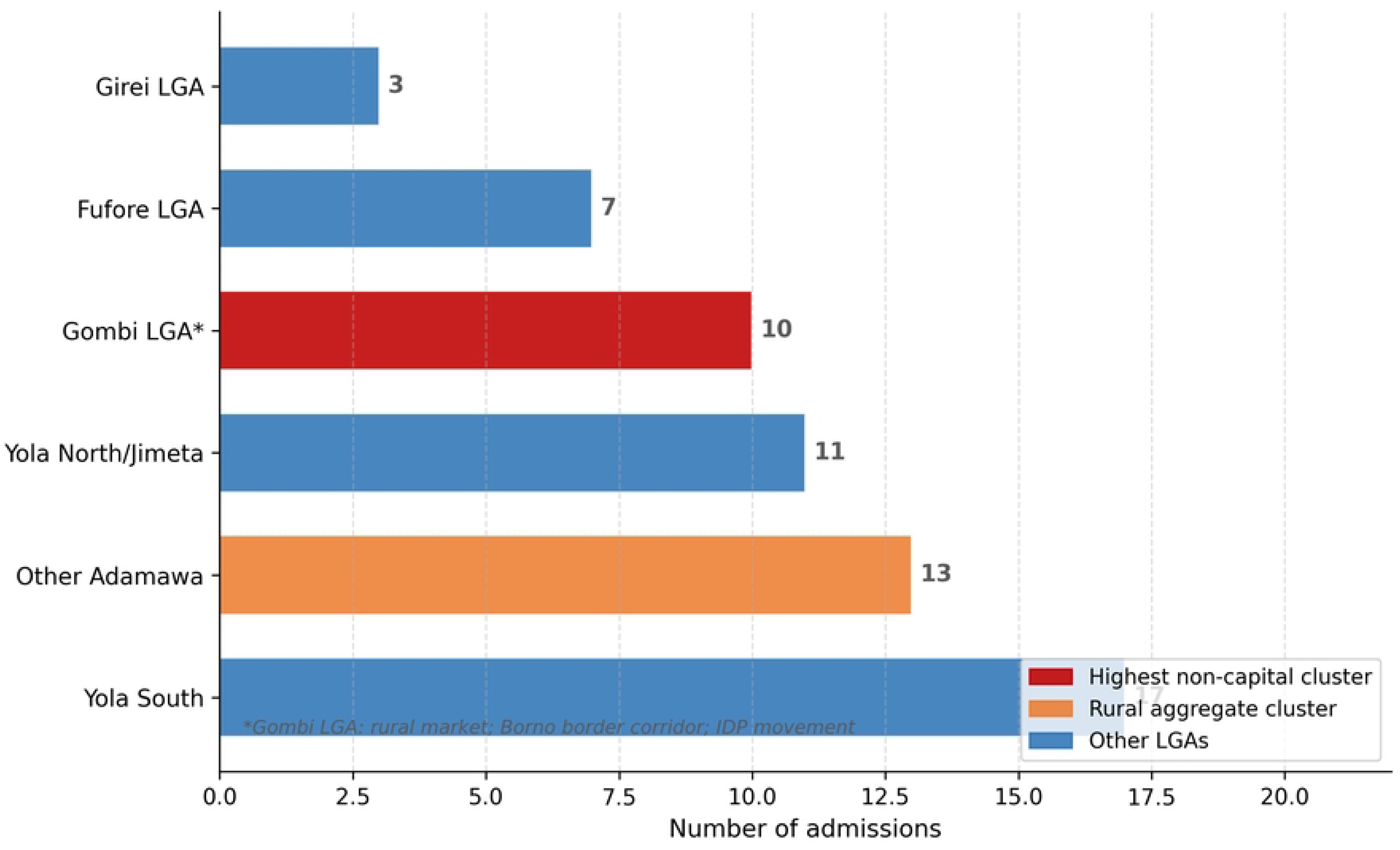
Geographic distribution of diphtheria admissions by local government area (LGA) of origin, MAUTH Yola, 2023–2026 (N=61). Gombi LGA (asterisk) was the highest-burden non-capital cluster, consistent with its proximity to Borno State’s outbreak epicentre and its role as a population movement corridor for displaced communities. LGA, local government area.

**Fig 6.**
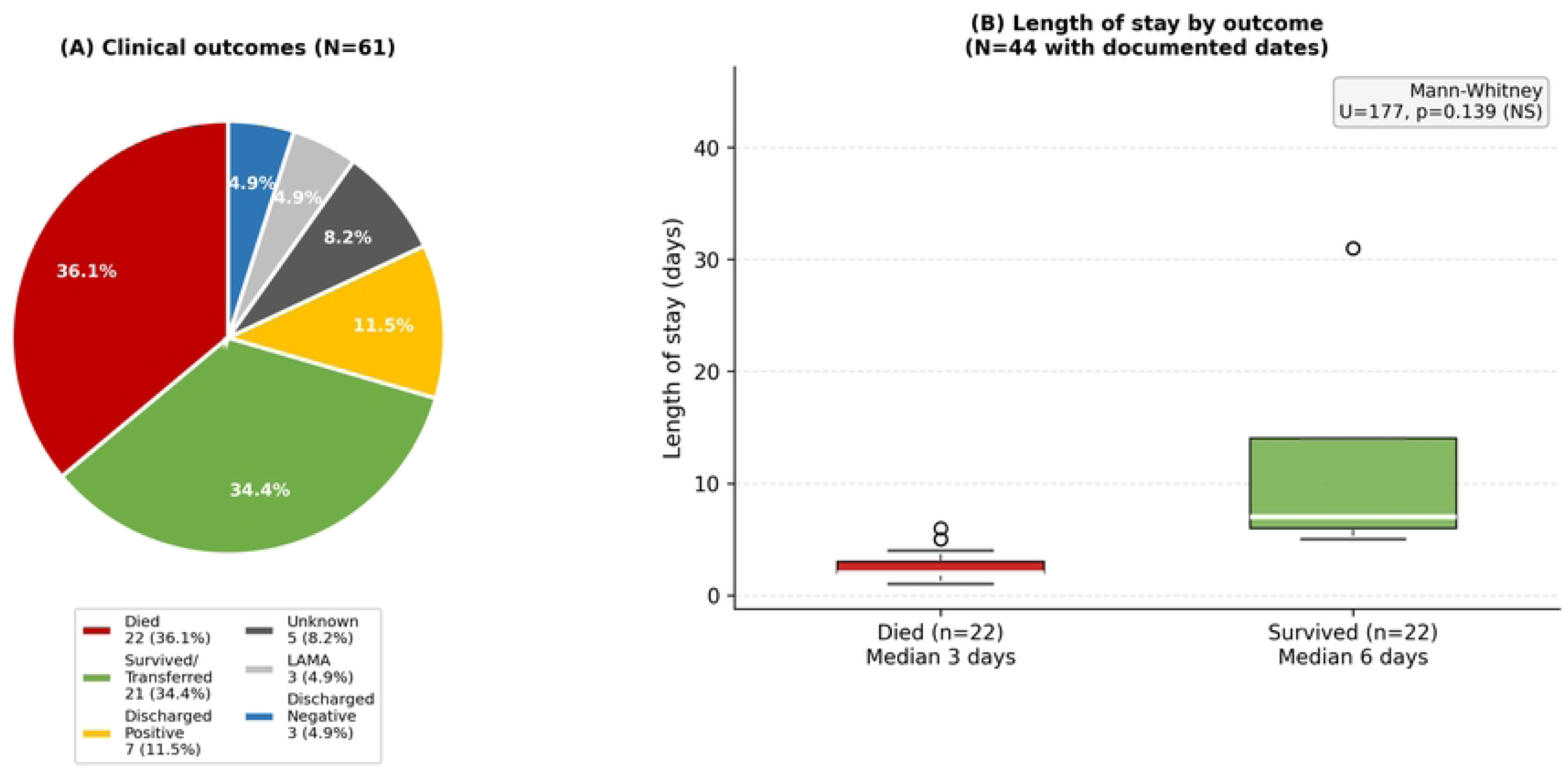
Clinical outcomes (panel A) and length of hospital stay by outcome (panel B), MAUTH Yola diphtheria admissions, 2023–2026. Panel A: outcome distribution (N=61). Panel B: length of stay by outcome (N=44 with documented dates). Patients who died had a shorter median length of stay (3 days) than survivors (6 days; Mann-Whitney U=177, p=0.139). LOS, length of stay; LAMA, left against medical advice.

## Discussion

### Summary of principal findings

This retrospective cohort study provides the first peer-reviewed, facility-level diphtheria epidemiological dataset from Adamawa State, north-eastern Nigeria. Sixty-one admissions over three and a half years showed a 580% escalation — from 5 cases in 2023 to 34 in 2025 — directly tracking Nigeria’s largest ever national epidemic. The overall in-hospital CFR of 41.5% substantially exceeds national surveillance averages, explained by referral bias, the complete absence of DAT during 2023, and the persistent shortage of IV erythromycin. The declining CFR (60.0% to 28.6%) against rising admission counts represents the most clinically important signal in the dataset — indicating that improving case management is reducing fatality even as community transmission accelerates.

### Contextualisation against published evidence

The MAUTH CFR far exceeds Nigeria’s national confirmed-case CFR of 5.7% [4]. It is higher than the 25.2% CFR documented at a dedicated north-western Nigeria isolation ward in 246 prospectively enrolled confirmed cases [16], most likely reflecting the more severe clinical spectrum reaching a tertiary referral centre and the 2023–2024 drug access deficit at MAUTH. A Yobe State prospective diphtheria treatment unit cohort — the only other published north-eastern Nigerian clinical dataset — documented similar drug availability challenges [15]. Both the MAUTH CFR decline (to 28.6% in 2025) and the Yobe cohort support the inference that when DAT and IV erythromycin become reliably available, outcomes improve substantially. The WHO benchmark for settings with adequate DAT and antibiotic access is a CFR of 5–10% [3]; the gap between this benchmark and MAUTH’s current performance quantifies the residual mortality attributable to modifiable supply constraints.

The 91.8% under-15 predominance mirrors national surveillance data showing 72–80% of confirmed cases aged 1–14 years [6] and is consistent with the accumulation of susceptible children who received incomplete DTP primary series during COVID-19 pandemic disruptions in 2020–2022. As of October 2023, 63.9% of confirmed cases nationally were documented as unvaccinated or only partially vaccinated [10]. UNICEF estimated that only 41.7% of Nigerian children under 15 were fully vaccinated against diphtheria as of 2023 [17], with Adamawa State substantially below this national average.

### The September–October seasonal peak: an Adamawa-specific signal

The September–October aggregated seasonal peak (47.5% of admissions) diverges from the January–April dry-season pattern reported nationally and for the Kano-centred outbreak. In Adamawa State, September–October corresponds to the post-harvest season, post-Eid al-Adha inter-community movement period, and the start of school reopening — conditions that combine population congregation with a large cohort of susceptible, unvaccinated children in enclosed environments. This distinct seasonal signal has direct public health implications: surveillance should be intensified and reactive vaccination campaigns pre-positioned before August each year in Adamawa State, not in January as might be inferred from the national seasonal profile.

### Gombi LGA: a priority transmission bridge

Gombi LGA’s emergence as the highest non-capital burden cluster (16.4% of cases) is a novel finding with direct policy implications. Gombi is a rural market town in northern Adamawa State on trade and migration routes connecting Adamawa to Borno State — the BAY-states zone contributing 271 of 417 BAY-state cases in early 2024 [9]. The geographic proximity to Borno’s outbreak epicentre, combined with documented IDP movement into northern Adamawa, strongly suggests that Gombi functions as a transmission bridge between the Borno epidemic and Adamawa State. We recommend that Gombi LGA be designated a targeted surveillance priority area by the Adamawa State Epidemiology Unit and the NCDC North-East Zonal Office.

### Diphtheria antitoxin shortage and the cost of administrative delay

The shortage of DAT and the delays inherent in its release pathway constitute the most consequential modifiable driver of mortality identified in this study. DAT supply in Nigeria has been constrained at every level throughout the current epidemic: globally, only a small number of manufacturers produce equine DAT, and concurrent large outbreaks in several world regions have depleted the international supply [22]; nationally, Nigeria does not routinely stock DAT, and although the NCDC and partners procured 10,050 vials during the 2023 emergency response, this quantity was insufficient for national demand and was prioritised toward the highest-burden north-western states [7,22]; subnationally, Adamawa State — lower in the supply-chain priority than Kano and Yobe — received delayed and incomplete allocations [12]. At the facility level, MAUTH held no independent DAT stock and was wholly dependent on the state and national release chain for every case.

Beyond absolute scarcity, the *structure* of the DAT release pathway independently generated life-threatening delay. Each requisition had to pass from the treating clinician through the Local Government Area and State DSNO and State Epidemiologist to the national Emergency Operations Centre for authorisation before release [7,21] — a process poorly suited to a disease in which antitoxin must be administered within hours of clinical suspicion. Published data indicate that diphtheria mortality rises steeply with treatment delay: fatality increases from approximately 4% among patients treated with antitoxin within 24–48 hours to 16.1% among those treated later [21], and a north-eastern Nigerian cohort documented a doubling of fatality with each 24-hour delay in appropriate treatment [15]. At MAUTH, these delays were most acute during weekends and on-call hours, when the surveillance-officer reporting chain and the national release mechanism operated with reduced capacity — precisely the periods when a child presenting with an obstructing pseudomembrane or evolving myocarditis can least afford to wait. The result was that patients with toxin-mediated disease sometimes deteriorated irretrievably before DAT could be sourced and given. Two structural reforms follow directly from this experience: first, the pre-positioning of a standing, quality-assured DAT buffer stock at MAUTH and other designated treatment centres in Adamawa State, held under a standing release authorisation that does not require case-by-case national approval during an active outbreak; and second, the establishment of a 24-hour, 7-day DAT release and DSNO escalation pathway so that weekend and out-of-hours presentations are not systematically disadvantaged. Together with reliable intravenous erythromycin supply, these measures would address the two most important modifiable contributors to the high case fatality documented in this study.

### Intravenous erythromycin shortage: a modifiable mortality driver

Our documentation of persistent IV erythromycin unavailability at MAUTH constitutes an important, previously unreported finding in the Nigerian diphtheria outbreak literature. The WHO Clinical Management of Diphtheria Guideline (2024) gives a strong recommendation for macrolides over penicillin specifically because Nigerian isolates show universal penicillin resistance [8]. The clinical cascade from IV erythromycin shortage is mechanistically coherent: inadequate antibiotic delivery through the only safe route in dysphagic patients allows ongoing bacterial toxin production, progressive pseudomembrane extension, and airway narrowing — directly explaining the 80% CFR in respiratory distress presentations. Oral azithromycin — a preferred alternative in the WHO and MSF guidelines for patients who can swallow — does not resolve the IV-route deficit for those who cannot [8,13]. Sustained pre-positioning of IV erythromycin (or IV azithromycin) as mandatory standing emergency stock at MAUTH, replenished before each August–October risk period, is the single most actionable clinical intervention identified in this study.

### Strengths and limitations

Strengths include: the first peer-reviewed, longitudinal, facility-level diphtheria dataset from Adamawa State; six full years of isolation ward records from the region’s only federal tertiary referral facility; application of Wilson score CIs for annual CFRs; and complication-stratified mortality analysis. Limitations include: retrospective paper-based register data with absent vaccination history, DAT and antibiotic timing, pseudomembrane extent, and severity scoring; predominantly clinical (rather than laboratory-confirmed) case ascertainment; limited statistical power (N=53 for regression); referral bias inflating the CFR; inability to calculate population-denominator incidence rates; and LOS data missing for 27.9% of admissions. A prospective multicentre study with electronic case report forms, laboratory confirmation, and standardised clinical severity scoring would substantially advance the evidence base for north-eastern Nigerian diphtheria management.

## Ethics statement

This study was approved by the Health Research Ethics Committee of Modibbo Adama University Teaching Hospital, Yola, Adamawa State, Nigeria (MAUTHYOLA/HREC/26/437), dated 26th March 2026. The committee waived the requirement for individual informed consent because the study used routinely collected, de-identified records. The work was conducted in accordance with the Declaration of Helsinki.

## Data availability

All data underlying the findings are contained within the manuscript and its Supporting Information. The de-identified, analysis-ready dataset (330 records) and a variable codebook are provided as S1 Dataset; the analysis code is available from the corresponding author on reasonable request.

## Author contributions

HA and AH conceived the study and abstracted data; HA and AH analysed the data; HA and AH interpreted results and drafted the manuscript; all authors approved the final version.

## Acknowledgments

The authors acknowledge the staff of the Isolation Ward and Medical Records Department of Modibbo Adama University Teaching Hospital, Yola, for their assistance with data collection and retrieval of historical ward records. The Nigeria Centre for Disease Control and Prevention (NCDC) is acknowledged for making diphtheria national situation reports publicly available, which provided essential comparator data for contextualising MAUTH findings within the national epidemic.

## Supporting information

**S1 Table. Logistic regression output.** Complete binary logistic regression results for predictors of in-hospital mortality among diphtheria admissions at MAUTH Yola, 2023–2026 (N=53 known outcomes). Includes regression coefficients (β), standard errors (SE), adjusted odds ratios, 95% confidence intervals, p-values, and model statistics (Nagelkerke pseudo-R²=0.127; −2LL=65.4). Reference: Female sex; <5 years age group.

**S2 Table. Length of hospital stay.** Length of hospital stay by outcome and age group among diphtheria admissions at MAUTH Yola, 2023–2026 (N=44 with documented dates). Includes mean ± SD, median, interquartile range, and range by clinical outcome and paediatric age group, with Mann-Whitney U test results (U=177, p=0.139).

**S3 Table. Geographic distribution.** Full geographic distribution of diphtheria admissions by local government area (LGA) of origin, MAUTH Yola, 2023–2026 (N=61), including case counts, deaths, case fatality rates, and contextual notes per LGA cluster.

**S4 Table. Comparative case fatality rate benchmarks.** Comparative case fatality rate benchmarks: MAUTH Yola 2023–2026 versus national Nigeria confirmed-case CFR (NCDC, 2025), BAY States CFR (WHO, 2024), NW Nigeria hospital cohort (Muhammad et al., BMC Infect Dis, 2025), Yobe State NE Nigeria cohort (Musa et al., Egypt J Intern Med, 2024), Borno State 2011 outbreak, and WHO global DAT-access benchmarks.

**S1 Fig. STROBE checklist.** STROBE (Strengthening the Reporting of Observational Studies in Epidemiology) checklist for cohort studies, completed for the MAUTH Yola diphtheria retrospective cohort study, 2023–2026.

**S2 Fig. Comparative CFR benchmark chart.** Bar chart comparing the MAUTH Yola overall CFR (41.5%) against published national, regional, and international diphtheria case fatality rate benchmarks. MAUTH entry highlighted in dark blue. CFR, case fatality rate.

**S1 Data. Anonymised summary dataset.** Anonymised case-level summary dataset (XLSX format) for all 61 diphtheria admissions at MAUTH Yola, 2023–2026. Fields: sequential case identifier; year of admission; month of admission; sex; age group; under-15 status; clinical presentation category; LGA cluster; outcome category; mortality indicator; length of stay (days). No patient names, exact dates of birth, or individual identifying information included. Ethics statement included in dataset documentation sheet.

